# Increased ^1^H-NMR metabolomics-based health score associates with declined cognitive performance and functional independence in older adults at risk of cardiovascular disease

**DOI:** 10.1101/2023.12.21.23300037

**Authors:** Michelle H. Zonneveld, Nour Al Kuhaili, Simon P. Mooijaart, P. Eline Slagboom, J. Wouter Jukema, Raymond Noordam, Stella Trompet

**Author notes:** **Corresponding author** Stella Trompet PhD, Department of Internal Medicine, Section of Gerontology and Geriatrics Leiden University Medical Center, PO Box 9600, 2300 RC Leiden, The Netherlands.

## Abstract

**Background:** The 1-HMR metabolomics-based MetaboHealth score, comprised of 14 serum metabolic markers, associates with disease-specific mortality, but it is unclear whether the score also reflects cognitive changes and functional impairment.

**Objectives:** Assess the associations between the MetaboHealth score with cognitive and daily functioning, and cognitive and functional decline in older people at increased cardiovascular risk.

**Methods:** 5292 older people free of dementia at baseline with mean age 75.3 years (standard deviation=3.4) from the Prospective Study of Pravastatin in the Elderly (PROSPER). MetaboHealth score was measured at baseline, and cognitive function and functional independence were measured at baseline and every 3 months during up to 2.5-years follow-up. Cognitive function was assessed using the Stroop test (selective attention), the Letter Digit Coding test (LDCT) (processing speed), and the two versions of the Picture Learning test (delayed and immediate; memory). Two tests of functional independence were used: Barthel Index (BI) and instrumental activities at daily living (IADL).

**Results:** A higher MetaboHealth score was associated with worse cognitive function (in all domains) and with worse functional independence. For example, after full adjustments, a 1-SD higher MetaboHealth score was associated with 9.02 (95%CI 7.29, 10.75) seconds slower performance on the Stroop test and 2.79 (-3.26,-2.21) less digits coded on the LDCT. During follow-up, 1-SD higher MetaboHealth score was associated with an additional decline of 0.53 (0.23,0.83) seconds on the Stroop test and -0.08 (-0.11,-0.06) points on the IADL.

**Conclusion:** Metabolic disturbance, as reflected by an increased metabolomics-based health score, may mark future cognitive and functional decline.

## 1. Introduction

In recent years, there has been growing interest in understanding the complex pathophysiology of ageing and its contribution to the development of disease. Ageing is a heterogenous process, highly variable between individuals, and often includes large discrepancies between physical and psychological functioning [1, 2]. As a result, there are large differences in the risk of age-related diseases between older individuals of the same calendar age. Recent research suggests that many chronic age-associated diseases like cognitive impairment, share “accelerated biological aging” as the underlying pathophysiological mechanism [3]. However, biological aging cannot be captured using only a single measurement.

In the last decade, attempts have been made to estimate biological age using blood-based molecular markers, such as DNA methylation and metabolomics assays, to construct health or biological age predictors such as the metaboAge score [4]. These molecular scores are based on the relation of molecules with chronological age, mortality, or biochemical risk factors [4]. However, these scores need to be tested in older individuals with regard to health outcomes and disease risk such as age-related cognitive impairment. Furthermore, investigating the underlying etiology between such molecular scores and outcomes like cognitive function will allow us to further understand pathophysiological pathways between risk-factors and outcomes.

Recently, Deelen et al. developed a score to predict disease-specific mortality using blood serum concentrations of 14 different metabolites (referred to as the “MetaboHealth score”), in a meta-analysis using data from 12 different cohort European-ancestry studies (comprising data of 44,168 individuals) [5]. Although the MetaboHealth score is an accurate predictor of various disease-specific outcomes and has been validated in various other populations [6], we were interested in understanding the pathway potentially leading to cognitive dysfunction and functional impairment. The association between the MetaboHealth score and cognitive function and daily functioning in older adults remains undetermined. Therefore, in this study we assessed the association between the MetaboHealth score, four domains of cognitive function, and functional independence in older people at higher risk of cardiovascular disease but free of baseline cognitive impairment.

## 2. Methods

### Study design and participants

Data from the Prospective Study of Pravastatin in the Elderly at Risk (PROSPER) was used for this study [7, 8]. PROSPER assessed the safety and efficacy of pravastatin in 5804 older individuals aged 70-82 years (from the Netherlands, Scotland, Ireland) with previous vascular disease or an increased risk of cardiovascular disease due to hypertension, smoking, and/or diabetes between December 1997 to May 1999. Participants were randomly assigned to receive either pravastatin 40 mg or placebo once daily, with an average treatment duration of 3.2 years. Both control as well as intervention group participants were included in the present study, as previous research showed pravastatin does not affect cognitive function [9]. Exclusion criteria for the participants included poor cognitive function (Mini-Mental State Examination <24 out of 30), current use of lipid-lowering drugs, current alcohol or drug abuse, and congestive heart failure (NYHA class III or IV). More details are described elsewhere [7, 8]. All participants who were included in the PROSPER study gave written informed consent. The study is in accordance with the Declaration of Helsinki and approved by the ethics review boards of the three centers in Leiden, Glasgow and Cork.

### Data collection

#### Metabolite measurements

Venous blood samples were collected from fasting participants at baseline before receiving the study medication and at 3-month intervals. Metabolites were measured at 6 months post-randomization, in 5329 participants. Both the placebo and statin groups were included as sensitivity analyses showed that there were no differences in outcomes when stratifying for treatment group.

Metabolite concentrations were measured using Nightingale proton nuclear magnetic resonance (^1^H-NMR) platform. This platform has been used widely in population-based studies of cardiometabolic diseases, and has been described in detail elsewhere [10–13]. After serum separation, the serum samples were stored at the biobank at -80°C. Plasma samples were mixed with an NMR buffer consisting of sodium phosphate. Hereafter, the low-molecular weight and lipoprotein metabolites data was collected using a spectrometer. To measure the lipid metabolite concentration and composition, a standardized lipid extraction was performed in the plasma samples. Various metabolites concentrations were measured, including lipid metabolites as VLDL-cholesterol, and low-molecular weight metabolites such as glutamine and creatinine [13].

The MetaboHealth score is based on the concentrations of: total lipids in chylomicrons and extremely large VLDL (XXL-VLDL-L); total lipids in small HDL (S-HDL-L); mean diameter for VLDL particles (VLDL-D); ratio of polyunsaturated fatty acids to total fatty acids (%, PUFA/FU); glucose; lactate; histidine; isoleucine; leucine; valine; phenylalanine; acetoacetate; albumin; and glycoprotein acetyls (GlycA). In the present study, participants were excluded if they had one or more missing measurements of the 14 metabolites. A higher score reflects a higher risk of mortality.

### Cognitive function and functional measurements

Cognitive function was measured every 3 months, with the time point of the last measurement varying from 36-48 months. The PROSPER study was conducted in three different countries and therefore colors, digits, and pictures were used instead of words [14]. A total of four neuropsychological tests were used to measure the changes in the cognitive function of the participants over time. The Stroop-Color-Word-Test (Stroop test) was used to assess selective attention [14]. The outcome variable of the Stroop test was the time in seconds needed to name or read the displayed 40 items. A higher Stroop score indicates a worse cognitive performance. Moreover, the minimally clinically important difference (MCID) of the Stroop test is 5.5 to 9.3 seconds [15]. To measure processing speed, the Letter-Digit Coding Test (LDCT) was used. The number of correctly entered digits in 60 seconds acts as the outcome variable [14]. The 15-Picture Word Learning test was used to assess immediate and delayed memory. Here, the outcome variable was the total number of the correctly recalled pictures, both immediately and after 20 minutes. For both versions of the Picture Learning Test as well as the Letter-Digit Coding Test, higher scores indicate better cognitive performance [14].

Functional status was assessed using the Barthel Index and IADL (Instrumental activities of daily living). The Barthel Index assesses self-care activities of daily living using 10 items, such as dressing, where a higher score indicates higher independence, with a maximum of 20 points [16]. Similarly, IADL measures activities of daily living using 7 items, but in addition includes the interaction with the social and physical environment [17]. Likewise, here a higher score also indicates higher functional capacity and independence.

### Covariates

Covariates were obtained during a 10-week screening period. These include demographic information (age, sex, and education), lifestyle (alcohol intake, smoking), and physical parameters (electrocardiogram, body mass index, blood pressure) [7]. The participant’s general practitioner reported on history of cardiovascular disease, diabetes mellitus, stroke, myocardial infarction, angina pectoris and transient ischemic attack. Hypertension was defined as a systolic blood pressure (SBP) ≥ 140 mmHg and/or diastolic blood pressure (DBP) ≥ 90 mmHg, or use of anti-hypertensives [18]. Diabetes mellitus was defined as a fasting blood glucose of ≥7 mmol/L.

### Statistical analyses

The baseline characteristics of the included participants are presented as means (with standard deviations) for continuous data and number (with percentages) for categorical data. The 14 MetaboHealth metabolites were log-transformed on a natural scale and standardized. Next, each metabolite was multiplied by their weight based on the meta-analyses results from Deelen *et al.* (ln(hazard ratio of all-cause mortality)) and summed for each included participant to calculate the individual MetaboHealth score [5].

The cross-sectional association between the MetaboHealth score and cognitive function performance was evaluated at 6 months using multivariable-adjusted linear regression model. Although metabolites were measured at 3 months after baseline, we chose to evaluate at 6 months as this time moment contained the largest sample size with complete cognitive function outcomes. The MetaboHealth score was analysed continuously (per SD) and in similarly-sized thirds, where the lower third was the reference category, indicating the lowest risk of mortality as reflected by the MetaboHealth score. Longitudinal associations between MetaboHealth score and repeated cognitive function measures during follow-up were assessed using linear mixed models.

Two different models were used to adjust for potential confounders. In the minimally adjusted model, we adjusted for sex, age, country, and education. The fully adjusted model additionally included history of cardiovascular disease, history of diabetes mellitus, history of myocardial infarction, smoking, alcohol intake, body mass index (BMI), systolic blood pressure, treatment arm, and apolipoprotein E (APOE ε4) genotype.

The results for the linear regression are reported as a beta-coefficient (β) for the cognitive test per SD increase in the MetaboHealth score, with a corresponding 95% confidence interval (95% CI). For the linear mixed models, the estimate of the additional decline for the interaction between time and the MetaboHealth score was reported with a corresponding 95% CI. SPSS Windows version 25 was used for all the analyses. Figures were designed using GraphPad Prism (version 9.3.1 for Windows, GraphPad Software, San Diego, California USA, www.graphpad.com).

### Sensitivity analyses

In addition to these overall analyses, we performed a number of sensitivity analyses to investigate the robustness of the associations considering different subgroups of the population. We performed separate analyses for placebo and pravastatin treatment groups, as well as stratifying for history of vascular disease. We also performed analyses excluding individuals who developed any of the following diseases during follow-up, to ensure a change in metabolomics did not follow as a result of the disease: non-fatal cancer; non-fatal stroke or transient ischemic attack; hospitalization due to heart failure; non-fatal coronary or cardiovascular events. Moreover, we analysed the association between individual metabolites included in the MetaboHealth score and cognitive decline, in order to see whether any of the individual metabolites were driving the overall observations done with the score. Individual metabolites were analysed continuously (per SD increase).

## 3. Results

The original PROSPER study included 5804 participants. Participants without metabolite measurements (n=475) or missing ≥1 of the 14 metabolites included in the MetaboHealth score (n=37) were excluded in the present study. A total of 5292 participants were included.

Table 1 displays the baseline characteristics of the study population. The mean age was 75.3 years (standard deviation (SD)=3.4), approximately half of the population was female (n=2733, 51.6%) and nearly half has a history of cardiovascular disease (n=2340, 44.2%). The mean body mass index (BMI) is 26.8 kg/m^2^ (SD=4.2) and a majority of the population has a history of hypertension (n=3280, 62.0%).

**Table 1.** Baseline characteristics of study population.

| **Sociodemographics** | **All (N=5292)** |
| --- | --- |
| Age, y, mean (SD) | 75.3 (3.4) |
| Female, n (%) | 2733 (51.6) |
| Age left school, y, mean (SD) | 15.1 (2.1) |
| Current smoker, n (%) | 1396 (26.4) |
| Alcohol intake, unit intake per week, mean (SD) | 5.2 (9.3) |
| **Cardiovascular risk factors** |  |
| History of CVD, n (%) | 2340 (44.2) |
| History of hypertension, n (%) | 3280 (62.0) |
| History of stroke or TIA, n (%) | 591 (11.2) |
| History of myocardial infarction, n (%) | 698 (13.2) |
| Systolic blood pressure, mmHg, mean (SD) | 154.7 (21.7) |
| Diastolic blood pressure, mmHg, mean (SD) | 83.8 (11.4) |
| Diabetes mellitus, n (%) | 561 (10.6) |
| Body mass index, kg/m<sup>2</sup>, mean (SD) | 26.8 (4.2) |
| Pravastatin treatment, n (%) | 2610 (49.3) |
| **MetaboHealth score metabolites** |  |
| MetaboHealth score, minimum; maximum | -1.25; 3.06 |
| MetaboHealth score, mean (SD) | -0.0009 (0.48) |
| Histidine, mmol/L, mean (SD) | 0.04 (0.01) |
| Leucine, mmol/L, mean (SD) | 0.06 (0.05) |
| Valine, mmol/L, mean (SD) | 0.14 (0.03) |
| Albumin, mmol/L, mean (SD) | 0.08 (0.01) |
| Glucose, mmol/L, mean (SD) | 3.23 (1.63) |
| Lactate, mmol/L, mean (SD) | 2.89 (1.65) |
| Isoleucine, mmol/L, mean (SD) | 0.05 (0.01) |
| Phenylalanine, mmol/L, mean (SD) | 0.05 (0.01) |
| Acetoacetate, mmol/L, mean (SD) | 0.06 (0.05) |
| Glycoprotein acetyls, mmol/L, mean (SD) | 1.29 (0.16) |
| Total lipids in chylomicrons and extremely large VLDL, mmol/L, mean (SD) | 0.03 (0.03) |
| Total lipids in small HDL, mmol/L, mean (SD) | 0.99 (0.09) |
| Mean diameter of VLDL-particles, nm, mean (SD) | 36.44 (1.19) |
| Ratio of polyunsaturated fatty acids to total fatty acids (PUFA/FU), mmol/L, mean (SD) | 36.09 (4.04) |
| **Cognitive function at month 9 of follow-up** |  |
| Stroop test, seconds, mean (SD)<sup>1</sup> | 64.5 (26.1) |
| LDCT, digits coded, mean (SD)<sup>2</sup> | 22.9 (7.8) |
| PLTi, pictures remembered, mean (SD)<sup>3</sup> | 9.5 (2.1) |
| PLTd, pictures remembered, mean (SD)<sup>3</sup> | 10.2 (2.8) |
| Barthel, index, mean (SD)<sup>4</sup> | 19.7 (0.9) |
| IADL, points, mean (SD)<sup>4</sup> | 13.5 (1.3) |

**Abbreviations:** CVD = cardiovascular disease; TIA = transient ischemic attack; VLDL = very low-density lipoprotein particle; HDL = high-density lipoprotein; SD = standard deviation; LDCT = Letter-Digit Coding Test; PLTi = Picture-Word Learning test immediate; PLTd = Picture-Word Learning Test delayed. <sup>1</sup>=performed by 3941 participants; <sup>2</sup>=performed by 4079 participants; <sup>3</sup>=performed by 4122 participants; <sup>4</sup>=performed by 5175 participants.

### Association between the MetaboHealth score and cognitive and daily function

Table 2 and Figure 1 show the cross-sectional associations between the MetaboHealth score and cognitive function. After full adjustments, in comparison to the lower third, the upper third of the MetaboHealth score – which indicates a higher health risk - was associated with a worse performance on all cognitive tests, but not the functional domain tests (Barthel index beta -0.02 (95% CI -0.09; 0.04)) and IADL -0.07 (95% CI -0.16; 0.02] points). To illustrate, the upper third coded 2.77 digits less (95%CI -3.30; -2.24) on the Letter-Digit coding test (LDCT) and performed 8.63 seconds slower (95% CI 6.68; 10.60) on the Stroop test, compared to the lower third. Similarly, compared to the lower tertile, the middle third of the MetaboHealth score was also associated with worse performance on three cognitive function tests, but was not associated with worse performance on the delayed version of the Picture-word learning test (PLTi) nor the functional tests.

**Figure 1.**
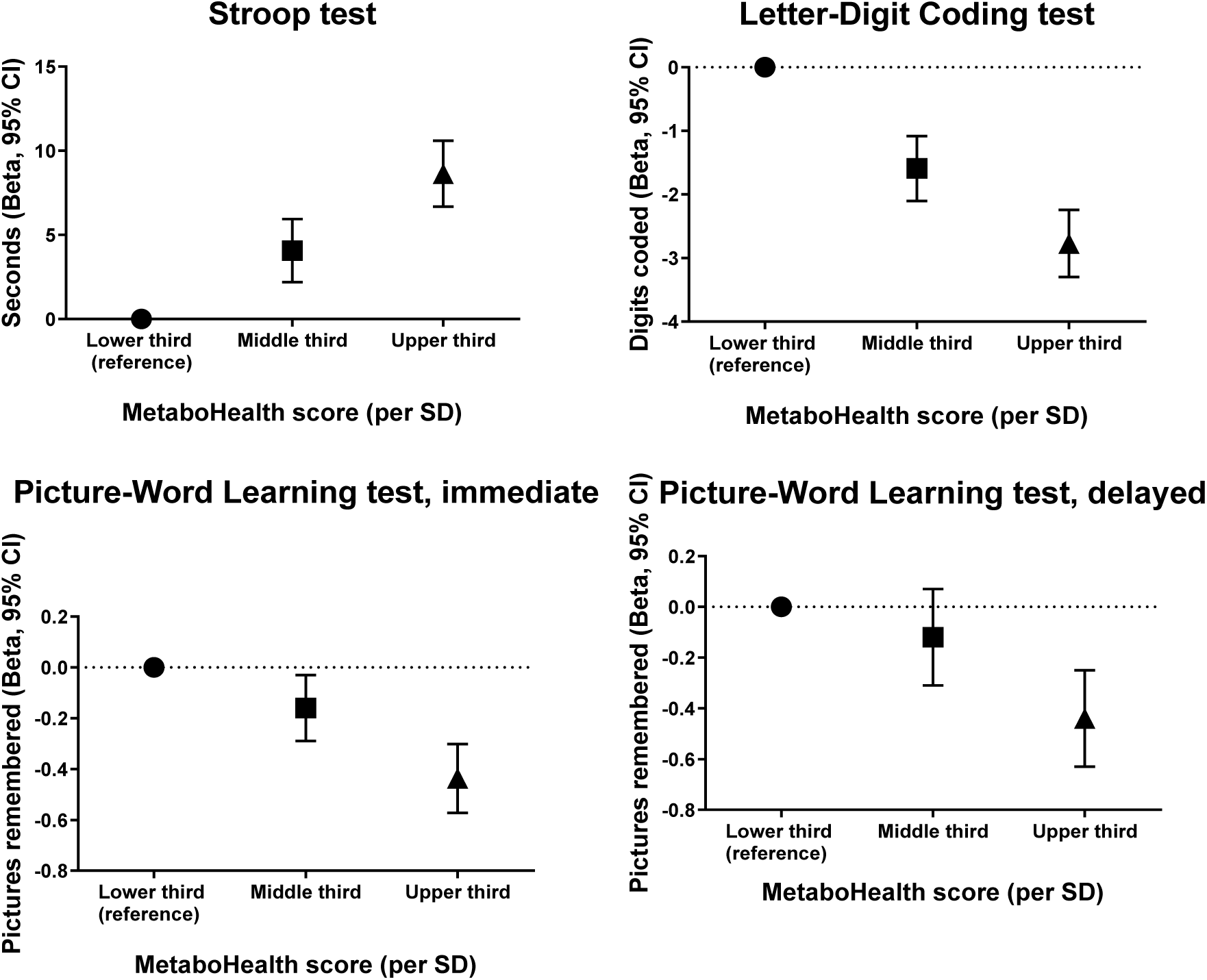
Fully adjusted cross-sectional associations of the MetaboHealth score (per SD) with cognitive function.

**Table 2.** Association between the MetaboHealth score and cognitive function.

| Cognitive test | MetaboHealth Score |  |  |  |
| --- | --- | --- | --- | --- |
|  | Lower third<br>(N<sub>max</sub>=1764)<br>Beta (95% CI) | Middle third<br>(N<sub>max</sub>=1764)<br>Beta (95% CI) | Upper third<br>(N<sub>max</sub>=1764)<br>Beta (95% CI) | Continuous (per SD)<br>(N<sub>max</sub>=5292)<br>Beta (95% CI) |
| **Minimally adjusted<sup>1</sup>** |  |  |  |  |
| Stroop, seconds | Ref | 3.98 (2.12; 5.85) | 9.20 (7.29; 11.12) | 9.51 (7.81; 11.2) |
| LDCT, digits coded | Ref | -1.57 (-2.07; -1.06) | -2.85 (-3.36; -2.33) | -2.89 (-3.35; -2.43) |
| PLTi, pictures remembered | Ref | -0.15 (-0.28; -0.03) | -0.45 (-0.58; -0.32) | -0.45 (-0.56; -0.33) |
| PLTd, pictures remembered | Ref | -0.12 (-0.30; 0.07) | -0.46 (-0.64; -0.27) | -0.53 (-0.69; -0.36) |
| Barthel, index | Ref | 0.06 (-0.01; 0.12) | -0.03 (-0.09; 0.03) | -0.15 (-0.20; -0.10) |
| IADL, points | Ref | 0.06 (-0.03; 0.14) | -0.09 (-0.18; 0.001) | -0.29 (-0.37; -0.22) |
| **Fully adjusted<sup>2</sup>** |  |  |  |  |
| Stroop, seconds | Ref | 4.07 (2.19; 5.94) | 8.63 (6.68; 10.60) | 9.02 (7.29; 10.75) |
| LDCT, digits coded | Ref | -1.59 (-2.10; -1.08) | -2.77 (-3.30; -2.24) | -2.79 (-3.26; -2.21) |
| PLTi, pictures remembered | Ref | -0.16 (-0.29; -0.03) | -0.44 (-0.57; -0.30) | -0.44 (-0.56; -0.32) |
| PLTd, pictures remembered | Ref | -0.12 (-0.31; 0.07) | -0.44 (-0.63; -0.25) | -0.51 (-0.68; -0.34) |
| Barthel, index | Ref | 0.06 (0.00; 0.13) | -0.02 (-0.09; 0.04) | -0.14 (-0.19; -0.09) |
| IADL, points | Ref | 0.06 (-0.03; 0.15) | -0.07 (-0.16; 0.02) | -0.28 (-0.35; -0.20) |

**Abbreviations:** LDCT = Letter-Digit Coding Test; PLTi = Picture-Word Learning test immediate; PLTd = Picture-Word Learning Test delayed; IADL = Instrumental activities of daily living. <sup>1</sup>=sex, age, country, education. <sup>2</sup>=sex, age, country, education, body mass index, history of cardiovascular disease, history of diabetes, history of myocardial infarct, smoking, alcohol intake, treatment arm, systolic blood pressure, apolipoprotein E genotype. MCID of the Stroop test: >5.5 seconds.

As a continuous variable, a higher MetaboHealth score was associated with lower scores in all cognitive and functional domains (all p<0.05). After full adjustments, a one SD higher MetaboHealth score was associated with 9.02 seconds slower (95% CI 7.29; 10.75) performance on the Stroop test, 2.79 less digits (95% CI -3.26; -2.21) coded on the LDCT, 0.44 less pictures remembered (95% CI -0.56; -0.32) on the immediate PLT and 0.51 less pictures remembered (95% CI 0.68; 0.34) on the delayed PLT. Similarly, a higher MetaboHealth score was also associated with a lower Barthel index (beta -0.14 points, 95% CI -0.19; -0.09) and lower IADL score (beta -0.28 points, 95% CI -0.35; -0.20).

### Association between the MetaboHealth score and cognitive and functional decline during follow-up

Table 3 displays the associations between the MetaboHealth score, cognitive decline and functional decline. A higher MetaboHealth score was associated with a faster decline in performance on all cognitive function tests and functional tests. After full adjustments, in comparison to the lower third, the upper third of the MetaboHealth score performed worse on all domains. For example, participants in the upper third showed an additional decline of 0.63 seconds (95% CI 0.30; 0.97) on the Stroop test, 0.11 less pictures remembered (95% CI -0.16; -0.07) on the PLTi and 0.07 points lower (95% CI -0.10; -0.04) on the IADL, compared to the lower third. A similar pattern was seen for the association between the middle third of the MetaboHealth score with the Stroop test and PLTd compared to the lower third, but not for the other cognitive and functional tests.

**Table 3.**
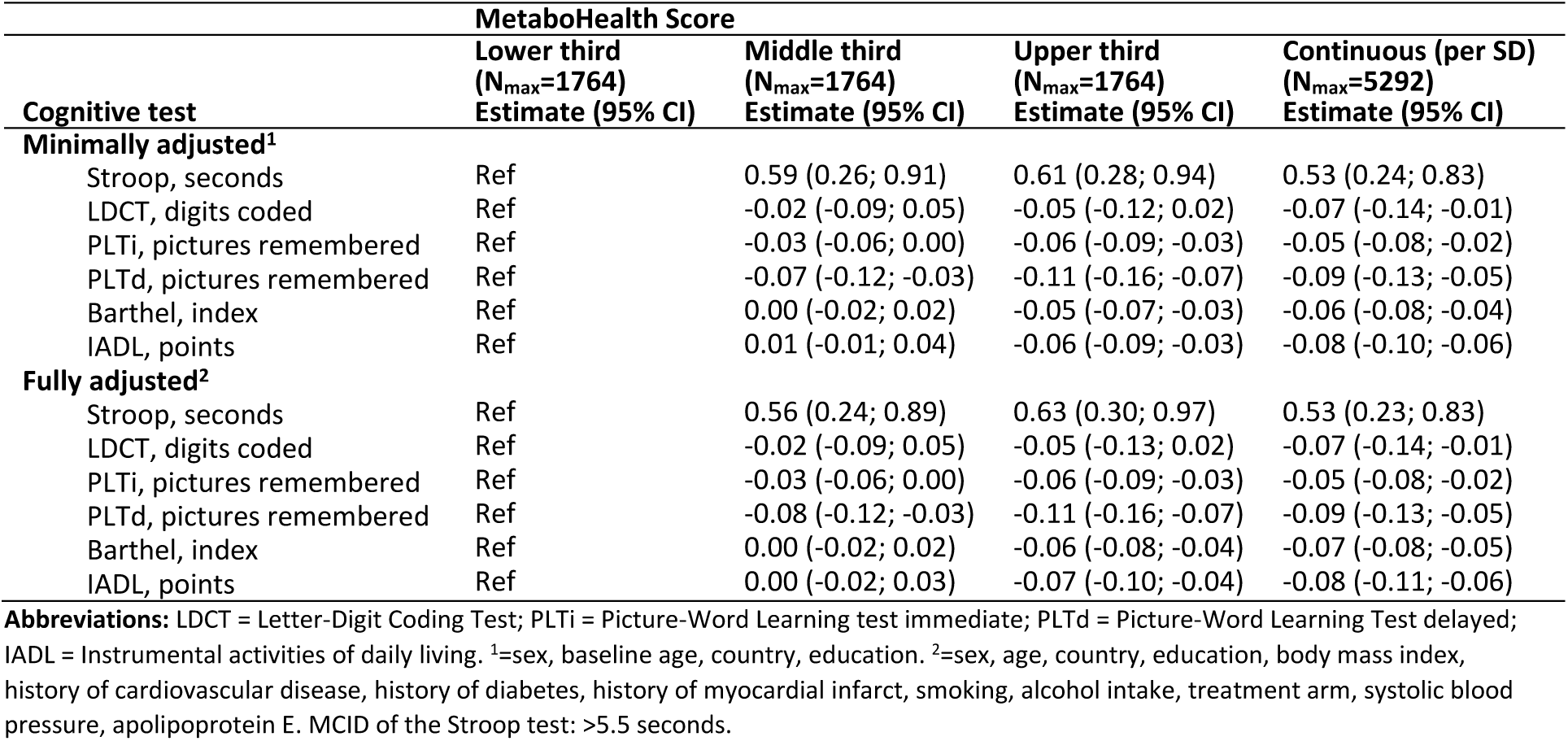
Association between the MetaboHealth score and cognitive decline.

Continuously, after full adjustments, a higher MetaboHealth score was associated with additional decline on all cognitive function tests and functional independence. For instance, a one SD higher MetaboHealth score was associated with an additional decline of 0.53 seconds (95% CI 0.23; 0.83) on the Stroop test, 0.07 digits coded (95% CI -0.14; -0.01) on the LDCT, -0.09 pictures remembered (95% CI -0.13; -0.05) on the PLTd, and -0.08 points (95% CI -0.11; -0.06) score on the IADL.

### Sensitivity analyses

The results were essentially unchanged when stratifying for the intervention and placebo group (Supplementary tables 1-2) and when stratifying for history of cardiovascular disease (Supplementary tables 3-4). Repeating the analyses after excluding participants with incident disease-states during follow-up also did not materially change the results (Supplementary tables 5-6).

Supplementary table 7 shows the association between the individual metabolites of the MetaboHealth score and cognitive decline. After full adjustments, various metabolites were associated with additional change in cognitive performance. To illustrate; higher concentrations of S-HDL-L, histidine and valine were associated with a better performance on the Stroop test during follow-up. For example, a one SD higher histidine was associated with an additional faster response of 0.24 seconds (95% CI -0.38; -0.10). Higher levels of glucose, acetoacetate and glycoprotein acetyls were associated with a lower score on the IADL. To illustrate; per SD higher glucose, there is an additional decline of -0.02 points on the IADL (95% CI -0.04; -0.01).

## 4. Discussion

In the present study, we investigated the association between the MetaboHealth score and cognitive function in a population of older adults at higher risk for cardiovascular disease, but free of dementia at baseline. Analyses showed a higher MetaboHealth score, based on 14 different serum metabolites, was associated with worse neurocognitive function and functional independence, both cross-sectionally and during follow-up. These findings were consistent in all tested cognitive and functional domains, and independent of cardiovascular risk-factors. Moreover, the beta-coefficients of the Stroop test overcame the minimal clinically important difference (MCID) (5.5; 9.3 seconds) as established by recent literature, indicating a clinically relevant change [15].

This is the first study describing the relationship between the MetaboHealth score, cognitive function and functional independence. Although the score has been validated as an accurate predictor of disease-related mortality, it has not yet been used to examine possible biological pathways leading to cognitive dysfunction and functional dependence. To our knowledge, studies examining metabolite-based scores and health outcomes like cognitive function and functional independence are scarce. Although recent literature describes relevant differences in various serum metabolite levels and health-related outcomes such as frailty [19], these studies cannot consistently account for the complex interactions between individual metabolites. The MetaboHealth score predicts various health-related types of mortality using 14 biomarkers involved in lipoprotein and fatty acid metabolism, inflammation, glycolysis, and fluid balance [5]. This has been validated in multiple study populations and has also been associated with frailty [5, 6]. In line, the metaboAge score has been recently developed and is able to predict biological age, also using metabolites [4]. Unlike the present study, only marginal associations were found between a higher metaboAge (indicating higher biological age) and cognitive function using data from the same study population [4]. This could be explained by the fact that the metaboAge score utilizes 56 metabolites compared to the 14 in the present study, and thus may be less sensitive to outliers or the association is more diluted [4, 5]. Although they do not associate with cognitive function in the same manner, both scores are strongly associated with vascular and all-cause mortality using various metabolites of which each is attributed a specific weight. Thus, the use of a score employing multiple metabolites such as the MetaboHealth or metaboAge score can be of added value for future studies investigating metabolites and ageing-related health outcomes.

Various large cohort studies have shown that lower levels valine and leucine, both branched-chain amino acids, are associated with a higher risk of dementia and AD [20–22], whereas elevated levels of glycoprotein acetyl have been associated with poorer cognitive performance [23]. The individual roles of metabolites may explain the difference in directionality, as glycoprotein acetyl is a marker of acute and systemic inflammation, and valine and leucine are primarily involved in metabolism [21]. Lipoprotein-associated phospholipase A_2_ (Lp-PLA_2_), a biomarker of vascular disorders in metabolic disease, has been identified as an independent risk-factor for cognitive impairment. Higher levels of Lp-PLA_2_ are associated with increased risk of Alzheimer’s disease and even more strongly with vascular dementia. This association was independent of BMI, blood pressure, blood glucose and blood lipids [24]. As found in the present study, the associations between the MetaboHealth score and cognitive function were more robust compared to the associations of individual metabolites. Although literature describes many relevant associations between single metabolites and cognition, these studies cannot account for the complex interactions between individual metabolites.

Several underlying pathways may justify the relationship between serum metabolites and cognitive function. The central nervous system (CNS) is highly enriched in lipids such as cholesterol, sphingolipids and polyunsaturated fatty acids (FA), which contribute to the structure of neuronal membranes and act as signaling molecules [25]. Neural networks employ a lipid-rich substance called myelin to protect nerve fibers from injury and propagate rapid signal transmission, which is mainly found in white matter. Previous studies have shown high numbers of white matter lesions in individuals with vascular cognitive impairment [26, 27]. [25]. In addition, a recent study found an increase in white matter hyperintensity volume to be associated with accelerated functional decline as measured by the Barthel index (-0.92 points per year, 95% CI -1.18; -0.67) [28]. The dry mass of white matter is mainly composed of proteins (15-30%) and lipids (70-85%), and thus, disturbances in the balance of lipids may be instrumental to (sub)clinical cognitive changes [25]. Similarly, phospholipids are also large constituents of pre- and postsynaptic membranes, and degeneration of neuronal membranes has been connected to synapse loss in Alzheimer’s Disease [21]. This could potentially also explain the results of the present study, as the MetaboHealth score includes four types of lipids.

In line, glutamine is widely described in literature in connection to the pathophysiology of dementia. Glutamine is an essential precursor for the biosynthesis of amino acid neurotransmitters such as glutamate, which is the major excitatory neurotransmitter in the CNS as well as the most abundant free amino acid in the brain [29]. Surprisingly, research has demonstrated that both higher glutamine and glutamate are associated with lower cognitive function [29], as high concentrations can potentially be harmful in a process called “excitotoxicity”. This may be explained by a disbalance in the cycle transferring glutamine from astrocytes to neurons and glutamate from neurons to astrocytes, as seen in Alzheimer’s disease [30]. Glycoprotein acetyls, also employed in the MetaboHealth score, have been found to predict 10-year mortality and are associated with lower general cognitive ability [21]. Increased concentrations of glycoprotein acetyls may also reflect systemic inflammation in various diseases such as cancer. Thus, this may in part explain the association with cognitive function as chronic diseases decrease cognitive performance.

This study has various strengths; the use of a large sample size of nearly 5000 older individuals as well as the use of multiple cognitive and functional outcome domains. Furthermore, we were able to demonstrate that the results are independent of numerous cardiovascular risk-factors. Limitations of this study include the relatively short follow-up period of 2.5-years. This is potentially insufficient time to fully reflect cognitive decline, although we did find significant associations. Moreover, the MCID’s of other cognitive tests have not yet been described in literature, preventing the direct translation of our results to a clinical setting. In line, the serum metabolites were measured only once during follow-up which may additionally not fully emulate underlying health changes. As cognitive decline is a neuropathological process of the CNS, it is important to consider the potential discrepancy between *peripheral* serum metabolites (as measured in the present study) and *central* metabolites, measurable in the cerebrospinal fluid or in brain tissue. The blood-brain barrier plays a key role in regulating the entrance and exit of metabolites into the CNS, and thus *central* changes may not always be reflected in peripheral blood. Nevertheless, we were still able to demonstrate relatively strong interactions between the MetaboHealth score and cognitive function in an older population at higher risk for cardiovascular disease.

In conclusion, a higher MetaboHealth score was associated with worse test performance and faster cognitive decline in multiple domains of cognitive functioning as well as higher functional dependence in older adults at higher risk of cardiovascular disease. The results are independent of cardiovascular risk-factors and development of incident disease states during follow-up. Moreover, the results of the Stroop test demonstrate a clinically relevant change by overcoming the MCID. Thus, metabolites and metabolite-based scores may act as risk-markers of future cognitive vulnerability and offer novel ways of identifying individuals at higher risk.

## Data Availability

All data produced in the present work are contained in the manuscript.

