## Supplementary tables for "Increased ^1^H-NMR metabolomics-based health score associates with declined cognitive performance and functional independence in older adults at risk of cardiovascular disease"

### Supplementary material.

**Supplementary table 1.** Association between the MetaboHealth score and cognitive function – stratified for treatment group.

| | **MetaboHealth Score (per SD)** | | | | | | |
|---|---|---|---|---|---|---|---|
| **Cognitive test** | **Lower third**<br>**Beta (95% CI)** | **Middle third Placebo**<br>**Beta (95% CI)** | **Middle third Pravastatin**<br>**Beta (95% CI)** | **Upper third Placebo**<br>**Beta (95% CI)** | **Upper third Pravastatin**<br>**Beta (95% CI)** | **Continuous Placebo**<br>**Beta (95% CI)** | **Continuous Pravastatin**<br>**Beta (95% CI)** |
| **Minimally adjusted[1]** | | | | | | | |
| Stroop, seconds | Ref | 4.60 (1.95; 7.24) | 3.27 (0.64; 5.89) | 8.32 (5.49; 11.14) | 10.09 (7.49; 12.69) | 9.47 (6.92; 12.02) | 9.75 (7.48; 12.03) |
| LDCT, digits coded | Ref | -1.71 (-2.40; -1.02) | -1.40 (-2.15; -0.65) | -2.36 (-3.09; -1.63) | -3.33 (-4.07; -2.59) | -2.64 (-3.30; -1.98) | -3.17 (-3.82;-2.53) |
| PLTi, pictures remembered | Ref | -0.12 (-0.30; 0.05) | -0.19 (-0.37; -0.01) | -0.44 (-0.63; -0.25) | -0.46 (-0.64; -0.28) | -0.45 (-0.61; -0.28) | -0.45 (-0.61; -0.30) |
| PLTd, pictures remembered | Ref | -0.14 (-0.40; 0.12) | -0.09 (-0.36; 0.17) | -0.53 (-0.80; -0.26) | -0.40 (-0.66; -0.14) | -0.58 (-0.82; -0.34) | -0.49 (-0.72; -0.27) |
| Barthel, index | Ref | 0.07 (-0.01; 0.15) | 0.04 (-0.05; 0.14) | 0.04 (-0.05; 0.12) | -0.10 (-0.19; 0.00) | -0.09 (-0.16; -0.02) | -0.20 (-0.28; -0.12) |
| IADL, points | Ref | 0.07 (-0.05; 0.19) | 0.04 (-0.09; 0.17) | 0.01 (-0.11; 0.14) | -0.18 (-0.31; -0.06) | -0.24 (-0.35; -0.14) | -0.34 (-0.45; -0.24) |
| **Fully adjusted[2]** | | | | | | | |
| Stroop, seconds | Ref | 4.62 (1.95; 7.28) | 3.48 (0.84; 6.13) | 7.37 (4.50; 10.24) | 9.80 (7.15; 12.46) | 8.68 (6.08; 11.27) | 9.38 (7.04; 11.71) |
| LDCT, digits coded | Ref | -1.68 (-2.38; -0.99) | -1.49 (-2.24; -0.74) | -2.18 (-2.93; -1.43) | -3.34 (-4.10; -2.58) | -2.44 (-3.21; -1.76) | -3.13 (-3.79; -2.47) |
| PLTi, pictures remembered | Ref | -0.14 (-0.32; 0.04) | -0.19 (-0.37; -0.01) | -0.43 (-0.63; -0.24) | -0.44 (-0.63; -0.26) | -0.43 (-0.61; -0.26) | -0.44 (-0.60; -0.28) |
| PLTd, pictures remembered | Ref | -0.14 (-0.40; 0.12) | -0.10 (-0.36; 0.17) | -0.49 (-0.77; -0.21) | -0.38 (-0.65; -0.11) | -0.55 (-0.80; -0.30) | -0.47 (-0.70; -0.24) |
| Barthel, index | Ref | 0.09 (0.01; 0.17) | 0.04 (-0.06; 0.13) | 0.06 (-0.03; 0.15) | -0.11 (-0.20; -0.01) | -0.07 (-0.15; -0.00) | -0.20 (-0.28; -0.12) |
| IADL, points | Ref | 0.09 (-0.03; 0.21) | 0.02 (-0.11; 0.15) | 0.04 (-0.09; 0.17) | -0.19 (-0.32; -0.06) | -0.22 (-0.33; -0.12) | -0.34 (-0.45; -0.23) |

**Abbreviations:** LDCT = Letter-Digit Coding Test; PLTi = Picture-Word Learning test immediate; PLTd = Picture-Word Learning Test delayed; IADL = Instrumental activities of daily living. [1]=sex, age, country, education. [2]=sex, age, country, education, body mass index, history of cardiovascular disease, history of diabetes, history of myocardial infarct, smoking, alcohol intake, systolic blood pressure, apolipoprotein E.

**Supplementary table 2.** Association between the MetaboHealth score and cognitive decline – stratified for treatment group.

| | **MetaboHealth Score (per SD)** | | | | | | |
|---|---|---|---|---|---|---|---|
| | **Lower third** | **Middle third Placebo** | **Middle third Pravastatin** | **Upper third Placebo** | **Upper third Pravastatin** | **Continuous Placebo** | **Continuous Pravastatin** |
| **Cognitive test** | **Estimate (95% CI)** | **Estimate (95% CI)** | **Estimate (95% CI)** | **Estimate (95% CI)** | **Estimate (95% CI)** | **Estimate (95% CI)** | **Estimate (95% CI)** |
| **Minimally adjusted[1]** | | | | | | | |
| Stroop, seconds | Ref | 0.58 (0.11; 1.05) | 0.59 (0.14; 1.04) | 0.65 (0.16; 1.14) | 0.57 (0.12; 1.02) | 0.42 (-0.03; 0.87) | 0.63 (0.23; 1.02) |
| LDCT, digits coded | Ref | -0.02 (-0.12; 0.08) | -0.02 (-0.12; 0.08) | -0.10 (-0.21; 0.00) | 0.00 (-0.10; 0.10) | -0.07 (-0.17; 0.02) | -0.07 (-0.16; 0.02) |
| PLTi, pictures remembered | Ref | -0.01 (-0.05; 0.03) | -0.04 (-0.08; 0.00) | -0.07 (-0.11; -0.02) | -0.06 (-0.10; -0.02) | -0.05 (-0.90; -0.01) | -0.05 (-0.08; -0.01) |
| PLTd, pictures remembered | Ref | -0.04 (-0.10; 0.02) | -0.11 (-0.17; -0.05) | -0.11 (-0.17; -0.05) | -0.12 (-0.18; -0.06) | -0.08 (-0.13; -0.02) | -0.11 (-0.16; -0.06) |
| Barthel, index | Ref | -0.01 (-0.04; 0.02) | 0.01 (-0.01; 0.04) | -0.06 (-0.09; -0.03) | -0.04 (-0.07; -0.02) | -0.06 (-0.09; -0.04) | -0.06 (-0.09; -0.04) |
| IADL, points | Ref | -0.01 (-0.05; 0.02) | 0.04 (0.00; -0.08) | -0.10 (-0.14; -0.06) | -0.02 (-0.06; 0.01) | -0.10 (-0.13; -0.06) | -0.07 (-0.10; -0.04) |
| **Fully adjusted[2]** | | | | | | | |
| Stroop, seconds | Ref | 0.49 (0.03; 0.96) | 0.64 (0.19; 1.10) | 0.60 (0.11; 1.09) | 2.63 (0.40; 4.87) | 0.36 (-0.08; 0.81) | 0.66 (0.26; 1.06) |
| LDCT, digits coded | Ref | -0.02 (0.12; 0.08) | -0.02 (-0.12; 0.08) | -0.11 (-0.21; 0.00) | -0.01 (-0.11; 0.10) | -0.07 (-0.17; 0.02) | -0.07 (-0.16; 0.02) |
| PLTi, pictures remembered | Ref | -0.02 (-0.06; 0.02) | -0.04 (-0.08; 0.00) | -0.07 (-0.11; -0.02) | -0.06 (-0.10; -0.02) | -0.05 (-0.09; -0.01) | -0.04 (-0.08; -0.01) |
| PLTd, pictures remembered | Ref | -0.04 (-0.10; 0.02) | 0.26 (0.03; 0.50) | -0.11 (-0.18; -0.05) | -0.12 (-0.18; -0.06) | -0.08 (-0.14; -0.02) | -0.11 (-0.16; -0.05) |
| Barthel, index | Ref | -0.02 (-0.05; 0.02) | 0.03 (-0.08; 0.13) | -0.07 (-0.10; -0.04) | -0.06 (-0.08; -0.02) | -0.07 (-0.10; -0.04) | -0.07 (-0.09; -0.04) |
| IADL, points | Ref | -0.02 (-0.06; 0.02) | 0.03 (-0.01; 0.07) | -0.10 (-0.14; -0.06) | -0.04 (-0.08; 0.00) | -0.10 (-0.13; -0.06) | -0.07 (-0.10; -0.04) |

**Abbreviations:** LDCT = Letter-Digit Coding Test; PLTi = Picture-Word Learning test immediate; PLTd = Picture-Word Learning Test delayed; IADL = Instrumental activities of daily living. [1]=sex, age, country, education. [2]=sex, age, country, education, body mass index, history of cardiovascular disease, history of diabetes, history of myocardial infarct, smoking, alcohol intake, systolic blood pressure, apolipoprotein E.

**Supplementary table 3.** Association between MetaboHealth score and cognitive function – stratified for history of cardiovascular disease.

| | MetaboHealth Score (per SD) | | | | | | |
|---|---|---|---|---|---|---|---|
| **Cognitive test** | **Lower third**<br>**Beta (95% CI)** | **Middle third**<br>**No**<br>**Beta (95% CI)** | **Middle third**<br>**Yes**<br>**Beta (95% CI)** | **Upper third**<br>**No**<br>**Beta (95% CI)** | **Upper third**<br>**Yes**<br>**Beta (95% CI)** | **Continuous**<br>**No**<br>**Beta (95% CI)** | **Continuous**<br>**Yes**<br>**Beta (95% CI)** |
| **Minimally adjusted[1]** | | | | | | | |
| Stroop, seconds | Ref | 4.45 (1.91; 7.00) | 3.37 (0.66; 6.08) | 9.65 (6.96; 12.34) | 8.69 (5.99; 11.40) | 8.84 (6.47; 11.21) | 10.45 (8.04; 12.86) |
| LDCT, digits coded | Ref | -1.56 (-2.24; -0.88) | -1.59 (-2.35; -0.83) | -2.79 (-3.50; -2.08) | -2.94 (-3.70; -2.18) | -2.78 (-3.40; -2.15) | -3.07 (-3.75; -2.39) |
| PLTi, pictures remembered | Ref | -0.14 (-0.30; 0.03) | -0.18 (-0.37; 0.02) | -0.40 (-0.58; -0.23) | -0.49 (-0.69; -0.30) | -0.42 (-0.57; -0.27) | -0.48 (-0.66; -0.31) |
| PLTd, pictures remembered | Ref | -0.15 (-0.39; 0.09) | -0.07 (-0.35; 0.22) | -0.40 (-0.65; -0.16) | -0.49 (-0.77; -0.20) | -0.50 (-0.72; -0.29) | -0.53 (-0.79; -0.28) |
| Barthel, index | Ref | 0.02 (-0.06; 0.10) | 0.10 (0.00; 0.21) | 0.03 (-0.04; 0.11) | -0.09 (-0.20; 0.01) | -0.05 (-0.11; 0.02) | -0.27 (-0.36; -0.18) |
| IADL, points | Ref | -0.01 (-0.10; 0.09) | 0.14 (-0.01; 0.30) | 0.01 (-0.10; 0.11) | -0.17 (-0.32; -0.01) | -0.13 (-0.21; -0.05) | -0.50 (-0.63; -0.36) |
| **Fully adjusted[2]** | | | | | | | |
| Stroop, seconds | Ref | 4.53 (1.97; 7.09) | 3.42 (0.67; 6.17) | 8.73 (6.02; 11.44) | 8.00 (5.22; 10.79) | 8.21 (5.82; 10.60) | 9.74 (7.24; 12.24) |
| LDCT, digits coded | Ref | -1.56 (-2.25; -0.87) | -1.63 (-2.39; -0.86) | -2.68 (-3.40; -1.95) | -2.79 (-3.57; -2.01) | -2.65 (-3.29; -2.01) | -2.89 (-3.59; -2.18) |
| PLTi, pictures remembered | Ref | -0.14 (-0.30; 0.03) | -0.19 (-0.39; 0.02) | -0.40 (-0.57; -0.22) | -0.46 (-0.67; -0.26) | -0.42 (-0.57; -0.26) | -0.45 (-0.63; -0.27) |
| PLTd, pictures remembered | Ref | -0.15 (-0.39; 0.09) | -0.06 (-0.35; 0.23) | -0.39 (-0.65; -0.14) | -0.42 (-0.71; -0.13) | -0.50 (-0.73; -0.28) | -0.47 (-0.73; -0.20) |
| Barthel, index | Ref | 0.03 (-0.04; 0.11) | 0.11 (0.00; 0.21) | 0.04 (-0.04; 0.12) | -0.09 (-0.19; 0.02) | -0.04 (-0.10; 0.03) | -0.27 (-0.36; -0.18) |
| IADL, points | Ref | 0.01 (-0.09; 0.10) | 0.13 (-0.03; 0.28) | 0.01 (-0.09; 0.11) | -0.16 (-0.32; 0.00) | -0.12 (-0.21; -0.04) | -0.48 (-0.61; -0.34) |

**Abbreviations:** LDCT = Letter-Digit Coding Test; PLTi = Picture-Word Learning test immediate; PLTd = Picture-Word Learning Test delayed; IADL = Instrumental activities of daily living. [1]=sex, age, country, education. [2]=sex, age, country, education, body mass index, history of diabetes, history of myocardial infarct, smoking, alcohol intake, treatment arm, systolic blood pressure, apolipoprotein E.

**Supplementary table 4.** Association between the MetaboHealth score and cognitive decline – stratified for history of cardiovascular disease.

| | **MetaboHealth Score (per SD)** | | | | | | |
|---|---|---|---|---|---|---|---|
| **Cognitive test** | **Lower third**<br>**Estimate (95% CI)** | **Middle third**<br>**No**<br>**Estimate (95% CI)** | **Middle third**<br>**Yes**<br>**Estimate (95% CI)** | **Upper third**<br>**No**<br>**Estimate (95% CI)** | **Upper third**<br>**Yes**<br>**Estimate (95% CI)** | **Continuous**<br>**No**<br>**Estimate (95% CI)** | **Continuous**<br>**Yes**<br>**Estimate (95% CI)** |
| **Minimally adjusted[1]** | | | | | | | |
| Stroop, seconds | Ref | 0.52 (0.08; 0.96) | 0.59 (-1.89; 3.07) | 0.56 (0.09; 1.02) | 0.69 (0.22; 1.16) | 0.55 (0.14; 0.96) | 0.52 (0.09; 0.95) |
| LDCT, digits coded | Ref | 0.03 (-0.06; 0.12) | -0.67 (-1.30; -0.04) | -0.09 (-0.19; 0.01) | 0.00 (-0.11; 0.10) | -0.13 (-0.21; 0.04) | 0.00 (-0.10; 0.10) |
| PLTi, pictures remembered | Ref | -0.03 (-0.07; 0.01) | -0.02 (-0.07; 0.02) | -0.06 (-0.10; -0.02) | -0.06 (-0.11; -0.02) | -0.06 (-0.09; -0.02) | -0.04 (-0.08; 0.00) |
| PLTd, pictures remembered | Ref | -0.05 (-0.10; 0.01) | -0.11 (-0.18; -0.04) | -0.13 (-0.19; -0.07) | -0.10 (-0.16; -0.03) | -0.12 (-0.17; -0.06) | -0.07 (-0.13; -0.01) |
| Barthel, index | Ref | 0.01 (-0.01; 0.04) | -0.01 (-0.05; 0.02) | -0.05 (-0.07; -0.02) | -0.06 (-0.10; -0.03) | -0.07 (-0.09; -0.04) | -0.06 (-0.09; -0.03) |
| IADL, points | Ref | 0.01 (-0.02; 0.05) | 0.01 (-0.03; 0.06) | -0.05 (-0.08; -0.01) | -0.07 (-0.11; -0.03) | -0.07 (-0.10; -0.04) | -0.08 (-0.12; -0.05) |
| **Fully adjusted[2]** | | | | | | | |
| Stroop, seconds | Ref | 0.48 (0.04; 0.92) | 0.69 (0.21; 1.16) | 0.53 (0.07; 1.00) | 0.77 (0.29; 1.25) | 0.51 (0.10; 0.92) | 0.56 (0.13; 1.00) |
| LDCT, digits coded | Ref | 0.04 (-0.06; 0.13) | -0.10 (-0.21; 0.01) | -0.08 (-0.18; 0.02) | -1.25 (-1.89; -0.62) | -0.12 (-0.21; -0.03) | 0.00 (-0.10; 0.09) |
| PLTi, pictures remembered | Ref | -0.03 (-0.07; 0.01) | -0.03 (-0.08; 0.01) | -0.06 (-0.19; -0.01) | -0.07 (-0.11; -0.02) | -0.06 (-0.09; -0.02) | -0.04 (-0.08; 0.00) |
| PLTd, pictures remembered | Ref | -0.05 (-0.10; 0.01) | -0.12 (-0.18; -0.05) | -0.13 (-0.19; -0.07) | -0.10 (-0.17; -0.03) | -0.11 (-0.17; -0.06) | -0.06 (-0.12; 0.00) |
| Barthel, index | Ref | 0.01 (-0.02; 0.03) | -0.01 (-0.05; 0.02) | -0.06 (-0.08; -0.03) | -0.06 (-0.10; -0.03) | -0.07 (-0.09; -0.04) | -0.06 (-0.09; -0.03) |
| IADL, points | Ref | 0.00 (-0.03; 0.03) | 0.02 (-0.03; 0.06) | -0.06 (-0.10; -0.03) | -0.07 (-0.12; -0.03) | -0.07 (-0.10; -0.05) | -0.09 (-0.13; -0.05) |

**Abbreviations:** LDCT = Letter-Digit Coding Test; PLTi = Picture-Word Learning test immediate; PLTd = Picture-Word Learning Test delayed; IADL = Instrumental activities of daily living. [1]=sex, age, country, education. [2]=sex, age, country, education, body mass index, history of diabetes, history of myocardial infarct, smoking, alcohol intake, treatment arm, systolic blood pressure, apolipoprotein E.

**Supplementary table 5.** Cross-sectional associations of MetaboHealth score and cognitive function – individuals with disease during follow-up excluded.

| | MetaboHealth Score (per SD) | | | |
|---|---|---|---|---|
| **Cognitive test** | **Lower third ($N_{max}$=1444) Beta (95% CI)** | **Middle third ($N_{max}$=1329) Beta (95% CI)** | **Upper third ($N_{max}$=1236) Beta (95% CI)** | **Continuous ($N_{max}$=4094) Beta (95% CI)** |
| **Minimally adjusted[1]** | | | | |
| Stroop, seconds | Ref | 5.05 (3.04; 7.07) | 9.11 (6.99; 11.22) | 8.33 (6.47; 10.20) |
| LDCT, digits coded | Ref | -1.93 (-2.50; -1.36) | -3.07 (-3.67; -2.47) | -2.97 (-3.50; -2.44) |
| PLTi, pictures remembered | Ref | -0.18 (-0.32; -0.04) | -0.47 (-0.62; -0.32) | -0.41 (-0.54; -0.28) |
| PLTd, pictures remembered | Ref | -0.20 (-0.41; 0.01) | -0.48 (-0.69; -0.26) | -0.50 (-0.70; -0.31) |
| Barthel, index | Ref | 0.01 (-0.05; -0.06) | -0.07 (-0.12; -0.01) | -0.08 (-0.14; -0.03) |
| IADL, points | Ref | -0.01 (-0.09; 0.08) | -0.14 (-0.23; -0.06) | -0.20 (-0.28; -0.12) |
| **Fully adjusted[2]** | | | | |
| Stroop, seconds | Ref | 4.89 (2.87; 6.91) | 8.42 (6.28; 10.56) | 7.85 (5.96; 9.74) |
| LDCT, digits coded | Ref | -1.86 (-2.44; -1.28) | -2.93 (-3.54;-2.32) | -2.87 (-3.42; -2.33) |
| PLTi, pictures remembered | Ref | -0.20 (-0.34; -0.05) | -0.47 (-0.63; -0.32) | -0.41 (-0.54; -0.27) |
| PLTd, pictures remembered | Ref | -0.22 (-0.43; -0.01) | -0.47 (-0.69; -0.25) | -0.50 (-0.70; -0.30) |
| Barthel, index | Ref | 0.02 (-0.03; 0.08) | -0.05 (-0.11; 0.01) | -0.07 (-0.12; -0.02) |
| IADL, points | Ref | 0.01 (-0.08; 0.09) | -0.11 (-0.20; -0.03) | -0.18 (-0.26; -0.10) |

**Abbreviations:** LDCT = Letter-Digit Coding Test; PLTi = Picture-Word Learning test immediate; PLTd = Picture-Word Learning Test delayed; IADL = Instrumental activities of daily living. [1]=sex, age, country, education. [2]=sex, age, country, education, body mass index, history of cardiovascular disease, history of diabetes, history of myocardial infarct, smoking, alcohol intake, treatment arm, systolic blood pressure, apolipoprotein E.

**Supplementary table 6.** Association between the MetaboHealth score and cognitive decline – individuals with disease during follow-up excluded.

| Cognitive test | MetaboHealth Score (per SD) Lower third ($N_{max}$=1444) Estimate (95% CI) | Middle third ($N_{max}$=1329) Estimate (95% CI) | Upper third ($N_{max}$=1236) Estimate (95% CI) | Continuous ($N_{max}$=4094) Estimate (95% CI) |
|---|---|---|---|---|
| **Minimally adjusted[1]** | | | | |
| Stroop, seconds | Ref | 0.58 (0.24; 0.91) | 0.58 (0.23; 0.93) | 0.59 (0.29; 0.90) |
| LDCT, digits coded | Ref | 0.00 (-0.08; 0.08) | -0.05 (-0.13; 0.03) | -0.08 (-0.15; -0.01) |
| PLTi, pictures remembered | Ref | -0.03 (-0.06; 0.00) | -0.06 (-0.09; -0.03) | -0.05 (-0.08; -0.02) |
| PLTd, pictures remembered | Ref | -0.06 (-0.10; -0.01) | -0.11 (-0.16; -0.06) | -0.09 (-0.13; -0.05) |
| Barthel, index | Ref | -0.01 (-0.02; 0.01) | -0.06 (-0.07; -0.04) | -0.04 (-0.06; -0.03) |
| IADL, points | Ref | 0.00 (-0.02; 0.03) | -0.05 (-0.08; -0.03) | -0.04 (-0.07; -0.02) |
| **Fully adjusted[2]** | | | | |
| Stroop, seconds | Ref | 0.58 (0.24; 0.92) | 0.61 (0.26; 0.97) | 0.59 (0.28; 0.91) |
| LDCT, digits coded | Ref | 0.00 (-0.07; 0.08) | -0.05 (-0.13; 0.03) | -0.08 (-0.16; -0.01) |
| PLTi, pictures remembered | Ref | -0.03 (-0.06; 0.00) | -0.06 (-0.09; -0.02) | -0.05 (-0.08; -0.02) |
| PLTd, pictures remembered | Ref | -0.05 (-0.10; 0.00) | -0.10 (-0.15; -0.05) | -0.09 (-0.13; -0.04) |
| Barthel, index | Ref | -0.01 (-0.03; 0.01) | -0.06 (-0.08; -0.04) | -0.05 (-0.06; -0.03) |
| IADL, points | Ref | 0.00 (-0.02; 0.02) | -0.05 (-0.08; -0.03) | -0.05 (-0.07; -0.02) |

**Abbreviations:** LDCT = Letter-Digit Coding Test; PLTi = Picture-Word Learning test immediate; PLTd = Picture-Word Learning Test delayed; IADL = Instrumental activities of daily living. [1]=sex, age, country, education. [2]=sex, age, country, education, body mass index, history of cardiovascular disease, history of diabetes, history of myocardial infarct, smoking, alcohol intake, treatment arm, systolic blood pressure, apolipoprotein E.

**Supplementary table 7.** Association between individual metabolites of the MetaboHealth score and cognitive decline.

| | Metabolites Z-score (per SD) | | | | | | | | | | | | | |
|---|---|---|---|---|---|---|---|---|---|---|---|---|---|---|
| | XXL-VLDL-L | S-HDL-L | VLDL-D | PUFA/FA | Glc | Lac | His | Ile | Leu | Val | Phe | AcAce | Alb | GlycA |
| **Cognitive test** | **Estimate (95% CI)** | **Estimate (95% CI)** | **Estimate (95% CI)** | **Estimate (95% CI)** | **Estimate (95% CI)** | **Estimate (95% CI)** | **Estimate (95% CI)** | **Estimate (95% CI)** | **Estimate (95% CI)** | **Estimate (95% CI)** | **Estimate (95% CI)** | **Estimate (95% CI)** | **Estimate (95% CI)** | **Estimate (95% CI)** |
| **Minimally adjusted[1]** | | | | | | | | | | | | | | |
| Stroop, seconds | -0.03 (-0.16; 0.11) | **-0.24 (-0.38; -0.10)** | -0.10 (-0.23; 0.04) | -0.08 (-0.22; 0.06) | 0.03 (-0.11; 0.17) | 0.00 (-0.14; 0.14) | **-0.23 (-0.37; -0.09)** | -0.05 (-0.19; 0.09) | -0.10 (-0.24; 0.04) | **-0.20 (-0.34; -0.06)** | -0.10 (-0.24; 0.04) | -0.05 (-0.20; 0.10) | -0.09 (-0.23; 0.04) | 0.03 (-0.11; 0.17) |
| LDCT, digits coded | -0.01 (-0.04; 0.02) | 002 (-0.01; 0.05) | 0.01 (-0.02; 0.04) | -0.01 (-0.04; 0.02) | -0.02 (-0.05; 0.01) | 0.02 (-0.01; 0.05) | 0.02 (-0.01; 0.05) | **0.04 (0.01; 0.07)** | **0.04 (0.01; 0.07)** | **0.05 (0.02; 0.08)** | **0.04 (0.01; 0.07)** | -0.02 (-0.05; 0.02) | **0.04 (0.01; 0.07)** | -0.01 (-0.04; 0.02) |
| PLTi, pictures remembered | -0.003 (-0.02; 0.01) | 0.00 (-0.01; 0.01) | 0.00 (-0.01; 0.01) | 0.02 (0.00; 0.03) | 0.00 (-0.01; 0.02) | -0.01 (-0.02; 0.00) | 0.01 (0.00; 0.02) | 0.00 (-0.01; 0.01) | 0.00 (-0.01; 0.01) | 0.01 (0.00; 0.02) | -0.01 (-0.02; 0.00) | 0.00 (-0.01; 0.01) | 0.00 (-0.01; 0.01) | -0.01 (-0.02; 0.00) |
| PLTd, pictures remembered | 0.01 (-0.003; 0.03) | 0.00 (-0.02; 0.02) | 0.02 (0.00; 0.04) | 0.02 (0.00; 0.04) | 0.01 (-0.01; 0.03) | -0.02 (-0.04; 0.00) | **0.03 (0.01; 0.04)** | **0.03 (0.01; 0.05)** | **0.03 (0.01; 0.05)** | **0.03 (0.02; 0.05)** | 0.00 (-0.02; 0.01) | -0.01 (-0.03; 0.01) | -0.01 (-0.03; 0.00) | -0.01 (-0.03; 0.01) |
| Barthel, index | **-0.01 (-0.02; -0.01)** | **0.01 (0.00; 0.02)** | 0.00 (-0.01; 0.01) | 0.00 (-0.01; 0.01) | -0.01 (-0.02; 0.00) | -0.01 (-0.02; 0.00) | 0.01 (0.00; 0.02) | 0.00 (-0.01; 0.00) | 0.01 (0.00; 0.02) | **0.02 (0.01; 0.02)** | 0.00 (-0.01; 0.01) | **-0.01 (-0.02; -0.004)** | 0.00 (-0.01; 0.01) | **-0.02 (-0.03; -0.01)** |
| IADL, points | -0.01 (-0.02; 0.00) | 0.00 (-0.01; 0.02) | 0.01 (0.00; 0.02) | 0.01 (0.00; 0.02) | **-0.02 (-0.03; -0.01)** | 0.01 (0.00; 0.02) | 0.02 (-0.01; 0.03) | 0.01 (0.00; 0.02) | **0.04 (0.02; 0.04)** | **0.03 (0.02; 0.04)** | 0.01 (0.00; 0.02) | **-0.02 (-0.03; -0.01)** | 0.00 (-0.01; 0.01) | **-0.02 (-0.03; -0.01)** |
| **Fully adjusted[2]** | | | | | | | | | | | | | | |
| Stroop, seconds | -0.03 (-0.17; 0.11) | **-0.24 (-0.38; -0.10)** | -0.10 (-0.24; 0.04) | -0.09 (-0.23; 0.05) | -0.001 (-0.15; 0.15) | 0.03 (-0.11; 0.17) | **-0.24 (-0.38; -0.10)** | -0.04 (-0.18; 0.10 | -0.10 (-0.24; 0.04) | **-0.19 (-0.33; -0.05)** | -0.11 (-0.25; 0.03) | -0.04 (-0.19; 0.10) | -0.09 (-0.23; 0.05) | 0.03 (-0.11; 0.17) |
| LDCT, digits coded | -0.01 (-0.04; 0.02) | 0.02 (-0.01; 0.05) | 0.01 (-0.02; 0.04) | -0.01 (-0.04; 0.02) | -0.02 (-0.05; 0.01) | 0.02 (-0.01; 0.06) | 0.02 (-0.01; 0.05) | **0.04 (0.01; 0.07)** | **0.04 (0.01; 0.07)** | **0.05 (0.02; 0.08)** | **0.04 (0.01; 0.07)** | -0.01 (-0.05; 0.02) | **0.04 (0.01; 0.07)** | -0.01 (-0.04; 0.02) |
| PLTi, pictures remembered | -0.003 (-0.02; 0.01) | 0.00 (-0.02; 0.01) | 0.00 (-0.02; 0.01) | 0.02 (0.01; 0.03) | 0.00 (-0.01; 0.01) | -0.01 (-0.02; 0.00) | 0.01 (0.00; 0.02) | 0.00 (-0.01; 0.01) | 0.00 (-0.01; 0.01) | 0.01 (0.00; 0.02) | -0.01 (-0.02; 0.00) | 0.00 (-0.01; 0.02) | 0.00 (-0.01; 0.01) | -0.01 (-0.02; 0.00) |
| PLTd, pictures remembered | **0.01 (0.00; 0.03)** | **0.01 (0.00; 0.03)** | 0.02 (0.00; 0.03) | 0.03 (0.01; 0.04) | 0.01 (0.01; 0.03) | -0.02 (-0.04; 0.00) | **0.02 (0.01; 0.04)** | **0.03 (0.01; 0.05)** | **0.03 (0.01; 0.05)** | **0.03 (0.02; 0.05)** | 0.00 (-0.02; 0.02) | -0.01 (-0.03; 0.01) | -0.01 (-0.03; 0.01) | -0.01 (-0.03; 0.01) |
| Barthel, index | -0.01 (-0.02; -0.01) | **0.01 (0.00; 0.02)** | 0.00 (-0.01; 0.01) | 0.00 (-0.01; 0.01) | -0.01 (-0.02; 0.00) | -0.01 (-0.02; 0.00) | 0.01 (0.00; 0.02) | 0.00 (-0.01; 0.01) | 0.01 (0.00; 0.02) | **0.02 (0.01; 0.02)** | 0.00 (-0.01; 0.01) | -0.01 (-0.02; 0.00) | 0.00 (-0.01; 0.01) | **-0.02 (-0.03; -0.01)** |
| IADL, points | -0.01 (-0.02; 0.00) | 0.01 (-0.01; 0.02) | 0.01 (0.00; 0.02) | 0.01 (-0.01; 0.02) | **-0.02 (-0.04; -0.01)** | 0.01 (0.00; 0.02) | **0.02 (0.01; 0.03)** | 0.01 (0.00; 0.02) | **0.03 (0.02; 0.04)** | **0.03 (0.02; 0.04)** | 0.01 (0.00; 0.02) | **-0.01 (-0.02; -0.002)** | 0.00 (-0.01; 0.02) | **-0.02 (-0.03; -0.01)** |

**Abbreviations:** XXL-VLDL-L = total lipids in chylomicrons and extremely large VLDL; S-HDL-L = total lipids in small HDL; VLDL-D = mean diameter for VLDL particles, PUFA/FA = ratio of polyunsaturated fatty acids to total fatty acids (%); Glc = glucose; Lac = lactate; His = histidine; Ile = isoleucine; Leu = leucine; Val = valine; Phe = phenylalanine; AcAce = acetoacetate; Alb = albumin; GlycA = glycoprotein acetyls; LDCT = Letter-Digit Coding Test; PLTi = Picture-Word Learning test immediate; PLTd = Picture-Word Learning Test delayed; IADL = Instrumental activities of daily living. [1]=sex, age, country, education. [2]=sex, age, country, education, body mass index, history of cardiovascular disease, history of diabetes, history of myocardial infarct, smoking, alcohol intake, treatment arm, systolic blood pressure, apolipoprotein E. Items written in bold indicate that the 95% confidence interval does not contain zero.
